# Delirium, Confusion, or Altered mental status as a risk for abnormal head computed tomogram findings in older adults in the emergency department: A systematic review and meta-analysis

**DOI:** 10.1101/2023.02.14.23285910

**Authors:** Sangil Lee, Faithe R. Cavalier, Jane M. Hayes, Michelle Doering, Alexander X. Lo, Danya Khoujah, Matthew A. Howard, Kerstin de Wit, Shan W. Liu

## Abstract

**Background:** Delirium, confusion, and altered mental status are common presentations among older adults to the emergency department (ED). We aimed to report the association between delirium, confusion or altered mental status in older ED patients and acute abnormal findings on head computed tomogram (CT).

**Methods:** A systematic review was conducted using Ovid Medline, Embase, Clinicaltrials.gov, Web of Science, and Cochrane Central from conception to April 8th, 2021. We included citations if they described patients aged 65 years or older who received head imaging at the time of ED assessment, and reported whether or not patients had delirium, confusion, or altered mental status. Screening, data extraction, and bias assessment were performed in duplicate. We estimated the odds ratios (OR) for abnormal neuroimaging in patients with altered mental status.

**Results:** The search strategy identified 3,014 unique citations, of which two studies reporting on 909 patients with delirium, confusion or altered mental status were included. No study formally assessed for delirium. The OR for abnormal head CT findings in patients with delirium, confusion or altered mental status was 0.35 (95% CI 0.031 to 3.97) compared to patients without delirium, confusion or altered mental status.

**Conclusion:** We did not find a statistically significant association between delirium, confusion or altered mental status and abnormal CT findings in older ED patients.

## INTRODUCTION

Delirium is a common emergency department (ED) presentation that increases hospital costs and is associated with higher mortality. Delirium is characterized by brain dysfunction such as confusion, altered level of consciousness, inattention, and perceptual disturbance.^1^ It is estimated that 6%-38% of older adults who present to the ED have delirium.^2–4^ Delirium presents a challenge in patient care as it is associated with higher mortality rates and functional decline. Studies estimated that delirium costs up to $152 billion US dollars each year in the healthcare setting.^1,5^ Consequently, many geriatric initiatives, including the multidisciplinary geriatric emergency department guidelines, focus on the screening, prevention, and treatment of delirium.^6^

The etiology of delirium is often multifactorial and includes infection, medications, pain, surgery, acute medical illness, drug intoxication, immobilization, metabolic derangement, sleep deprivation, and acute neurological disease.^7–13^ The neurological etiologies for delirium include intracranial hemorrhage, ischemic stroke, and brain tumor. Many of these neurological etiologies can be diagnosed with Computed Tomography (CT) or Magnetic Resonance Imaging (MRI) imaging. Head CT is commonly used in the ED to evaluate patients presenting with concern for a neurological emergency..^13,14^ However, there is variability in the practice of obtaining head CT for older adult patients presenting with confusion, delirium or altered mental status.^15–17^ There is no consensus on whether delirium in older ED patients necessitates neuroimaging in all cases.^18,19^ We recently showed an incidence of 15% for abnormal neuroimaging in older ED patients with confusion or altered mental status.^20^ However, the analysis did not include a comparison group of older patients without delirium.

### Objective

We undertook a systematic review and meta-analysis to evaluate whether delirium in older ED patients increases the odds of finding abnormal pathology on neuroimaging compared to older ED patients without delirium. Given that many studies would include patients who present to the ED in an altered mental state and without a formal diagnosis of “delirium”, we expanded our review to capture the more encompassing description of “altered mental status”

## METHODS

### Overview

This manuscript was written in accordance with the Preferred Reporting Items for Systematic Reviews and Meta-Analyses (PRISMA) guidelines.^21^

### Search Strategy and Study Eligibility

We searched Ovid Medline from 1946, Embase from 1947, Web of Science from 1900, Cochrane Central, PubMed Central, and Clinicaltrials.gov up to April 8, 2021. The search strategies were created in collaboration with a medical librarian and can be viewed in Appendix A. The search strategies involved using many combinations of common key words and terms, such as (delirium OR confusion OR acute mental status change) and (computed tomography OR magnetic resonance imaging AND emergency department OR acute care OR emergency physician), filtered to exclude pediatric studies. The unique citations were stored in Covidence systematic review software, Veritas Health Innovation, Melbourne, Australia. Specifically for delirium, in order to maximize the sensitivity of our search parameters, we included the more generic terms to capture any instance of potential delirium by adding the terms “confusion” and ‘altered mental status”

We included original studies reporting on patients aged 65 years of age or older who had received neuroimaging during their ED visit. Included studies had to report on patients with and without delirium. We included confusion or altered mental status as a proxy measure for delirium because there is a overlap between these terms. We included patients diagnosed with delirium superimposed on dementia. We excluded conference abstracts and published manuscripts. Studies on hospitalized inpatients or specific subgroups of delirious patients (such as trauma patients or patients with cancer), were excluded. We also excluded case reports. Two team members independently screened the title and abstract of each citation based on the inclusion and exclusion criteria, and a third team member acted as the tiebreaker if it was needed. Then all team members were included in full-text reviews to ensure that each citation was independently reviewed by two team members, with a third acting as a tiebreaker if needed, to determine its final inclusion in this systematic review.

### Data Extraction and Quality Assessment

Data were extracted from each study by three team members using a spreadsheet template designed by the investigators. Disagreements were resolved by discussion. The template included information about the study author, years of conduct, research design, and sample size; population characteristics (age, sex); and abnormal neuroimaging findings (number and diagnosis). If the results of a study were reported in a manner that did not allow for accurate or complete data extraction, the authors were contacted to obtain primary data for the purpose of this meta-analysis.

Using the “Quality Assessment of Diagnostic Accuracy Studies-2 (QADAS-2)”,^22^ two team members independently checked for the risk of bias in each of the studies with a third team member acting as a tiebreaker if needed. ^23^ The tool assesses the representativeness of the general population, accuracy of the assessment of the outcome, and completeness of the data to categorize studies as having a low, intermediate, or high risk of bias.

### Statistical Analysis

We performed a meta-analysis of the collected data to calculate the pooled random effects estimated odds ratio (OR) and 95% confidence interval of abnormal neuroimaging in those who had delirium/confusion/altered mental status compared to those without delirium/confusion/altered mental status. Heterogeneity was appraised using I^2^. The meta-analysis was completed using MedCalc Statistical Software v20.019 (MedCalc Software Ltd, Ostend, Belgium).

## RESULTS

### Study Characteristics

The employed search strategy resulted in the identification of 3,035 studies. After title and abstract screnning, 3,031 were excluded because they did not meet the inclusion criteria or there was a duplicate. Forty-seven studies underwent a full text review, 45 were excluded and 2 studies included in the analysis. ^24,25^ Detail of the selection process can be found in Figure 1.

**Figure 1.**
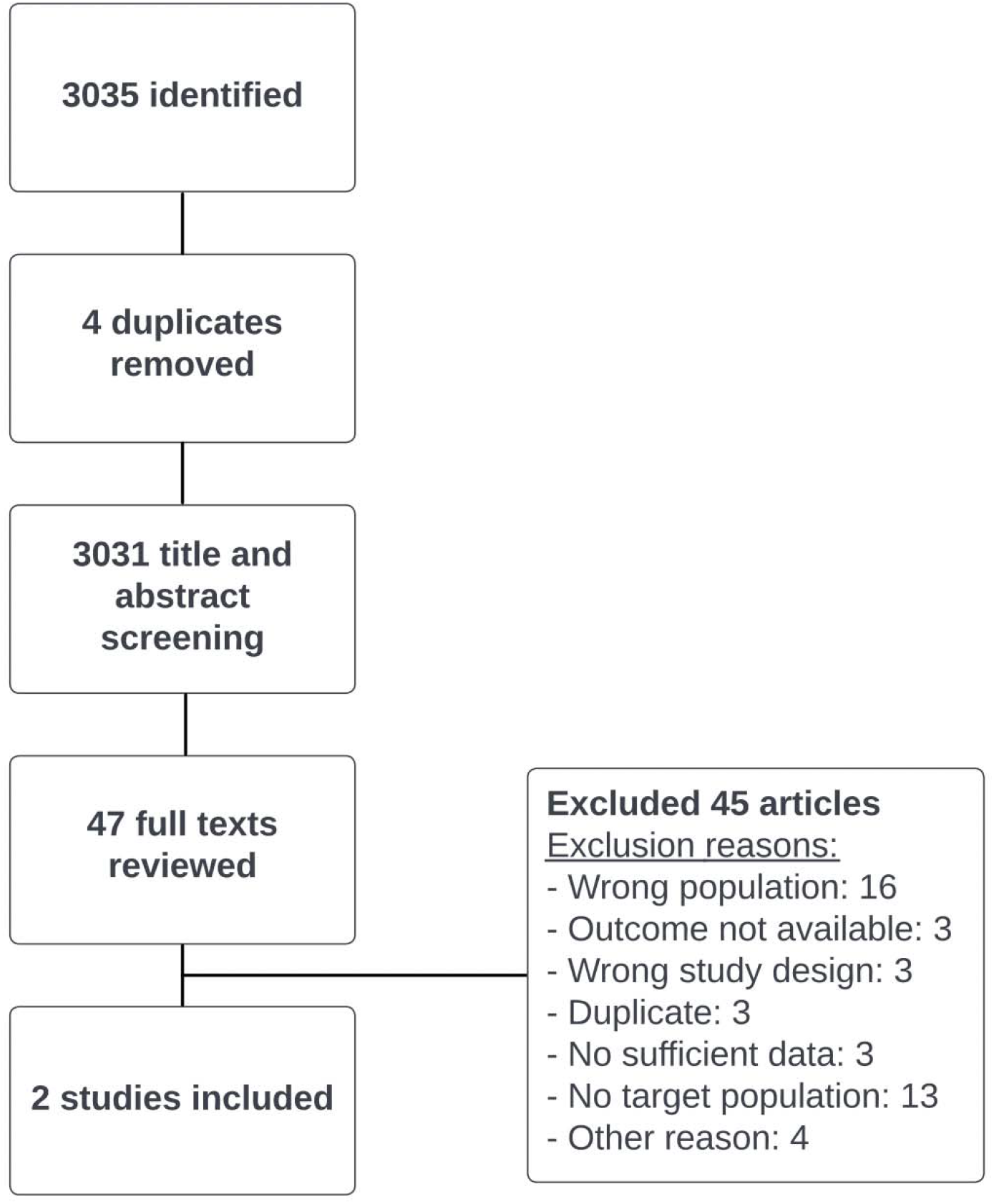
Flow chart showing article selection.

Both included studies used head CT imaging (not MRI).^24,25^ Both studies were retrospective cohort studies. We reported on a subset of the study by Nesselroth et al. who were 65 years and older.^24^ We considered patients described as “confused” to have delirium, confusion or altered mental status. Further, the authors categorized the abnormal CT results into “acute” or “chronic” and defined these terms.^24^ In the study by Segard et al., all patients were 75 years or older, and abnormal CT findings were described using the term “abnormal”, but a further definition was not clear.^25^ For the purposes of this analysis, we considered those classified as having ‘consciouness disorder with GCS < 14’ and ‘delirium without focal neurological deficits’ to be positive for delirium, confusion or altered mental status. ^25^ Table 1 shows more detailed characteristics of these studies.

**Table 1.** Characteristics of included studies.

| **Author**<br>**Country**<br>**Year** | **Study Design** | **Definition of Delirium** | **Study Period** | **Age** | **Sex** | **Included in the meta-analysis** | **N** | **Acute Abnormal Head CT n (%)** | **CT abnormality** |
| --- | --- | --- | --- | --- | --- | --- | --- | --- | --- |
| Nesselroth<br>Israel<br>2021 | Retrospective | Confusion | Jan 2017-Feb 2017 | Not reported | Not reported | Patients $\geq 65$ who had head CT in the ED | 86 | 17 (19.8%) | Not reported |
| Segard<br>France<br>2013 | Retrospective | GCS $< 14$ or delirium with no focal deficits | Not reported | Not reported | Not reported | Patients $\geq 75$ who had head CT in the ED | 103 | 41 (39.8%) | *112 Ischemic stroke<br>35 ICH<br>5 Brain mass<br>52 Other |

SD: Standard Deviation, CT: Computed Tomography, IQR: Inter Quartile Ratio, ICH: intracranial hemorrhage
\* some patients had more than one abnormality

### Risk of Bias

Nesselroth et al.^24^ showed low risk of bias for patient selection and reference test domains, high risk for index test, and unclear risk of bias for flow and timing domain. Segard et al.^25^ showed high risk of bias for index test and reference test and unclear risk of bias for patient selection and flow and timing domains. (Table 2.)

**Table 2.**
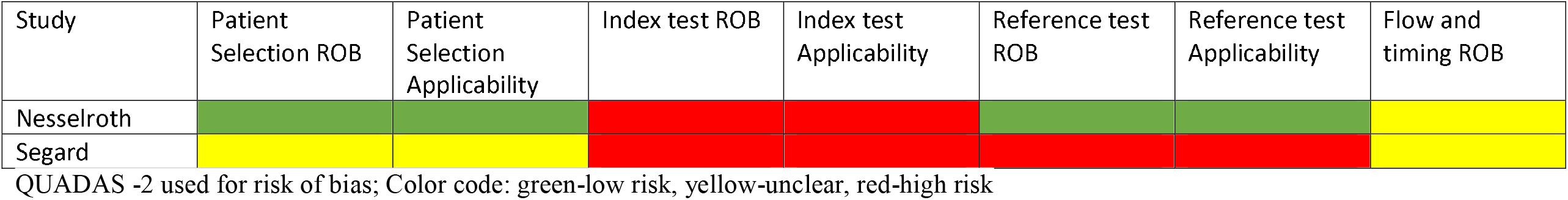
Risk of bias and concern for applicability for included study.

### Synthesis of Results

The pooled odds of patients with delirium having an abnormal neuroimaging compared to those without delirium was 0.35 (95% CI 0.031 to 3.97) and I^2^ was 97.25% with a p < 0.0001 indicating substantial heterogeneity. The pooled rate of abnormal neuroimaging was 30.7%. Figure 2 demonstrates the corresponding forest plot.

**Figure 2.**
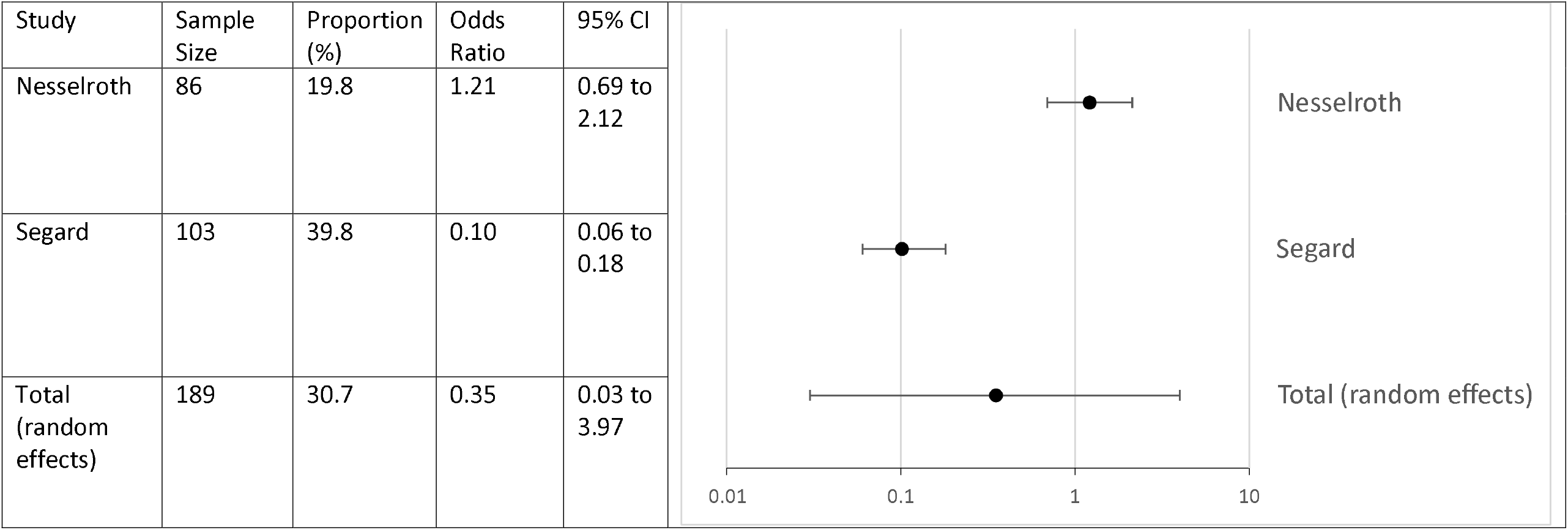
Forest plot showing a pooled odds ratios for abnormal head CT when altered mental status is present.

## DISCUSSION

This systematic review and meta-analysis showed that there is very limited quality research addressing whether delirium in older adults is associated with acute intracranial abnormality on head imaging in the ED. The existing research is insufficient to provide a conclusion. Given this limited quality of research, our meta-analysis could not provide conclusive evidence on whether delirium predicted abnormal acute findings on neuroimaging.

This is the first analysis comparing the risk of intracranial pathology among older ED patients with delirium to those who do not have delirium. Our systematic review had important findings. There were only two studies. None of the reviewed studies assessed all included patients for delirium, therefore misclassification bias is likely. Neither study used a validated delirium screening tool. Both studies used surrogate measures for delirium such as Glasgow Coma Scale less than 14 and confusion. Most importantly, the studies had unclear or high risk of bias, meaning that pooling of the results has little clinical meaning.

We previously showed that the rate of abnormal head CT was about 15% for older ED patients with altered mental status.^20^ Similarly, Akhtar et al. conducted a systematic review of the yield of head CT for patients with delirium or altered mental status, and reported that the overall percentage of head CT was 13% (95% CI: 10.2%–15.9%) in the inpatient/ED and 17.4% (95% CI: 10%–26.3%) in the ICU, with considerable heterogeneity.^26^ Importantly, Akhtar et al. evaluated delirium and altered mental status as a separate entities. ^26^ We suspect the higher rate of abnormal neuroimaging in our present study might be due to random error. Our present meta-analysis was based on a limited number of patients and studies, likely prone to selection bias. The number of abnormal head neuroimaging in these previous studies was not negligible, but our meta-analysis did not show sufficient evidence to support or refute the need for neuroimaging when a clinical feature of acute mental status change was present.

### Limitations

There are several limitations to this study. First, our study used a broader definition of delirium which included confusion and altered mental status because of the paucity of literature focusing on patients with delirium in the ED. However, the chief complaint of altered mental status is highly specific for delirium.^27^ The designation of altered mental status is not very sensitive to the presence of delirium, so we suspect that there were patients with delirium, particularly hypoactive delirium, who were classified as not having delirium, confusion or altered mental status in our analysis. Second, our work focused on older adults with delirium, confusion or altered mental status who received a head CT in the ED rather than all older patients in the ED. It is likely that the decision to order the head CT introduced spectrum bias. The true proportion of head CT abnormalities in patients with and without delirium may differ. Third, we found only two retrospective studies with high risk of bias.

## CONCLUSION

This meta-analysis showed that exisiting evidence is too poor to determine whether the presence of delirium, confusion or altered mental status is associated with abnormal neuroimaging findings in the ED.

## Data Availability

All data produced in the present study are available upon reasonable request to the authors

## Notes

### Competing Interest Statement

The authors have declared no competing interest.

### Author Declarations

Publicly available data.

## REFERENCES

1. Han JH, Schnelle JF, Ely EW. The relationship between a chief complaint of “altered mental status” and delirium in older emergency department patients. Acad Emerg Med. 2014;21(8):937–940. doi:10.1111/acem.12436

2. Han JH, Vasilevskis EE, Schnelle JF, et al. The diagnostic performance of the richmond agitation sedation scale for detecting delirium in older emergency department patients. Acad Emerg Med. 2015;22(7):878–882. doi:10.1111/acem.12706

3. Kennedy M, Enander RA, Tadiri SP, Wolfe RE, Shapiro NI, Marcantonio ER. Delirium risk prediction, healthcare use and mortality of elderly adults in the emergency department. J Am Geriatr Soc. 2014;62(3):462–469. doi:10.1111/jgs.12692

4. Carpenter CR, Hammouda N, Linton EA, et al. Delirium prevention, detection, and treatment in emergency medicine settings: A geriatric emergency care applied research (GEAR) network scoping review and consensus statement. Acad Emerg Med. 2021;28(1):19–35. doi:10.1111/acem.14166

5. Leslie DL, Marcantonio ER, Zhang Y, Leo-Summers L, Inouye SK. One-year health care costs associated with delirium in the elderly population. Arch Intern Med. 2008;168(1):27–32. doi:10.1001/archinternmed.2007.4

6. Carpenter CR, Bromley M, Caterino JM, et al. Optimal older adult emergency care: introducing multidisciplinary geriatric emergency department guidelines from the American College of Emergency Physicians, American Geriatrics Society, Emergency Nurses Association, and Society for Academic Emergency Medicine. Acad Emerg Med. 2014;21(7):806–809. doi:10.1111/acem.12415

7. Young J, Murthy L, Westby M, Akunne A, O’Mahony R, Guideline Development Group. Diagnosis, prevention, and management of delirium: summary of NICE guidance. BMJ. 2010;341:c3704. doi:10.1136/bmj.c3704

8. Wong J, Lam D, Choi S, et al. The prevention of delirium in elderly with obstructive sleep apnea (PODESA) study: protocol for a multi-centre prospective randomized, controlled trial. BMC Anesthesiol. 2018;18(1):1. doi:10.1186/s12871-017-0465-5

9. Wong A, Young AT, Liang AS, Gonzales R, Douglas VC, Hadley D. Development and Validation of an Electronic Health Record-Based Machine Learning Model to Estimate Delirium Risk in Newly Hospitalized Patients Without Known Cognitive Impairment. JAMA Netw Open. 2018;1(4):e181018. doi:10.1001/jamanetworkopen.2018.1018

10. Vasilevskis EE, Han JH, Hughes CG, Ely EW. Epidemiology and risk factors for delirium across hospital settings. Best Pract Res Clin Anaesthesiol. 2012;26(3):277–287. doi:10.1016/j.bpa.2012.07.003

11. Marcantonio ER, Goldman L, Orav EJ, Cook EF, Lee TH. The association of intraoperative factors with the development of postoperative delirium. Am J Med. 1998;105(5):380–384. doi:10.1016/s0002-9343(98)00292-7

12. Inouye SK, Zhang Y, Jones RN, Kiely DK, Yang F, Marcantonio ER. Risk factors for delirium at discharge: development and validation of a predictive model. Arch Intern Med. 2007;167(13):1406–1413. doi:10.1001/archinte.167.13.1406

13. Fong TG, Tulebaev SR, Inouye SK. Delirium in elderly adults: diagnosis, prevention and treatment. Nat Rev Neurol. 2009;5(4):210–220. doi:10.1038/nrneurol.2009.24

14. Sheng AZ, Shen Q, Cordato D, Zhang YY, Yin Chan DK. Delirium within three days of stroke in a cohort of elderly patients. J Am Geriatr Soc. 2006;54(8):1192–1198. doi:10.1111/j.1532-5415.2006.00806.x

15. Shobeirian F, Ghomi Z, Soleimani R, Mirshahi R, Sanei Taheri M. Overuse of brain CT scan for evaluating mild head trauma in adults. Emerg Radiol. 2021;28(2):251–257. doi:10.1007/s10140-020-01846-6

16. Cellina M, Panzeri M, Floridi C, Martinenghi CMA, Clesceri G, Oliva G. Overuse of computed tomography for minor head injury in young patients: an analysis of promoting factors. Radiol Med. 2018;123(7):507–514. doi:10.1007/s11547-018-0871-x

17. Naughton BJ, Moran M, Ghaly Y, Michalakes C. Computed tomography scanning and delirium in elder patients. Acad Emerg Med. 1997;4(12):1107–1110. doi:10.1111/j.1553-2712.1997.tb03690.x

18. Hardy JE, Brennan N. Computerized tomography of the brain for elderly patients presenting to the emergency department with acute confusion. Emerg Med Australas. 2008;20(5):420–424. doi:10.1111/j.1742-6723.2008.01118.x

19. Shenvi C, Kennedy M, Austin CA, Wilson MP, Gerardi M, Schneider S. Managing delirium and agitation in the older emergency department patient: the ADEPT tool. Ann Emerg Med. 2020;75(2):136–145. doi:10.1016/j.annemergmed.2019.07.023

20. Liu SW, Lee S, Hayes JM, et al. Head computed tomography findings in geriatric emergency department patients with delirium, altered mental status, and confusion: A systematic review. Acad Emerg Med. Published online November 4, 2022. doi:10.1111/acem.14622

21. Liberati A, Altman DG, Tetzlaff J, et al. The PRISMA statement for reporting systematic reviews and meta-analyses of studies that evaluate health care interventions: explanation and elaboration. J Clin Epidemiol. 2009;62(10):1–34. doi:10.1016/j.jclinepi.2009.06.006

22. Whiting PF, Rutjes AWS, Westwood ME, et al. QUADAS-2: a revised tool for the quality assessment of diagnostic accuracy studies. Ann Intern Med. 2011;155(8):529–536. doi:10.7326/0003-4819-155-8-201110180-00009

23. The CLARITY Group at McMaster University. Tool to Assess Risk of Bias in Longitudinal Symptom Research Studies Aimed at the General Popu;ation. Accessed December 23, 2022. https://www.distillersr.com/wp-content/uploads/2021/03/Tool-to-Assess-Risk-of-Bias-Longitudinal-Symptom-Research-Studies-Aimed-at-the-General-Population-DistillerSR.pdf

24. Nesselroth D, Klang E, Soffer S, et al. Yield of head CT for acute findings in patients presenting to the emergency department. Clin Imaging. 2021;73:1–5. doi:10.1016/j.clinimag.2020.11.025

25. Segard J, Montassier E, Trewick D, Le Conte P, Guillon B, Berrut G. Urgent computed tomography brain scan for elderly patients: can we improve its diagnostic yield? Eur J Emerg Med. 2013;20(1):51–53. doi:10.1097/MEJ.0b013e32834f9d51

26. Akhtar H, Chaudhry SH, Bortolussi-Courval É, et al. Diagnostic yield of CT head in delirium and altered mental status-A systematic review and meta-analysis. J Am Geriatr Soc. Published online November 26, 2022. doi:10.1111/jgs.18134

27. Han JH, Wilber ST. Altered mental status in older patients in the emergency department. Clin Geriatr Med. 2013;29(1):101–136. doi:10.1016/j.cger.2012.09.005

